# Transcriptional Signatures of Hippocampal Tau Pathology in Primary Age-Related Tauopathy and Alzheimer’s Disease

**DOI:** 10.1101/2023.09.12.23295440

**Authors:** Genevieve L Stein-O’Brien, Ryan Palaganas, Ernest M. Meyer, Javier Redding-Ochoa, Olga Pletnikova, Haidan Guo, William R Bell, Juan C Troncoso, Richard L Huganir, Meaghan Morris

**Affiliations:** McKusick-Nathans Institute of Genetic Medicine, Johns Hopkins School of Medicine, Baltimore, MD, USA; Single Cell Training and Analysis Center, Johns Hopkins University School of Medicine, Baltimore, MD, USA; Department of Neuroscience, Johns Hopkins University School of Medicine, Baltimore, MD, USA; Kavli Neuroscience Discovery Institute, Baltimore, MD; UPMC Hillman Cancer Center Cytometry Facility, University of Pittsburgh Medical Center, Pittsburgh, PA, USA; Department of Pathology, Johns Hopkins University School of Medicine, Baltimore, MD, USA; Department of Pathology and Anatomical Sciences, University at Buffalo, Buffalo, NY; Department of Pathology and Laboratory Medicine, Indiana University School of Medicine, Indianapolis, IN, USA; Department of Neurology, Johns Hopkins University School of Medicine, Baltimore, MD

## Abstract

**Background:** Tau pathology is common in age-related neurodegenerative diseases. Tau pathology in primary age-related tauopathy (PART) and in Alzheimer’s disease (AD) has a similar biochemical structure and anatomic distribution, which is distinct from tau pathology in other diseases. However, the molecular changes associated with intraneuronal tau pathology in PART and AD, and whether these changes are similar in the two diseases, is largely unexplored.

**Methods:** Using GeoMx spatial transcriptomics, mRNA was quantified in CA1 pyramidal neurons with tau pathology and adjacent neurons without tau pathology in 6 cases of PART and 6 cases of AD, and compared to 4 control cases without pathology. Transcriptional changes were analyzed for differential gene expression and for coordinated patterns of gene expression associated with both disease state and intraneuronal tau pathology.

**Results:** Synaptic gene changes and two novel gene expression signatures associated with intraneuronal tau were identified in PART and AD. Overall, gene expression changes associated with intraneuronal tau pathology were similar in PART and AD. Synaptic gene expression was decreased overall in neurons in AD and PART compared to control cases. However, this decrease was largely driven by neurons lacking tau pathology. Synaptic gene expression was increased in tau-positive neurons compared to tau-negative neurons in disease. Two novel gene expression signatures associated with intraneuronal tau were identified by examining coordinated patterns of gene expression. Genes in the up-regulated expression pattern were enriched in calcium regulation and synaptic function pathways, specifically in synaptic exocytosis. These synaptic gene changes and intraneuronal tau expression signatures were confirmed in a published transcriptional dataset of cortical neurons with tau pathology in AD.

**Conclusions:** PART and AD show similar transcriptional changes associated with intraneuronal tau pathology in CA1 pyramidal neurons, raising the possibility of a mechanistic relationship between the tau pathology in the two diseases. Intraneuronal tau pathology was also associated with increased expression of genes associated with synaptic function and calcium regulation compared to tau-negative disease neurons. The findings highlight the power of molecular analysis stratified by pathology in neurodegenerative disease and provide novel insight into common molecular pathways associated with intraneuronal tau in PART and AD.

**Graphical Abstract:** 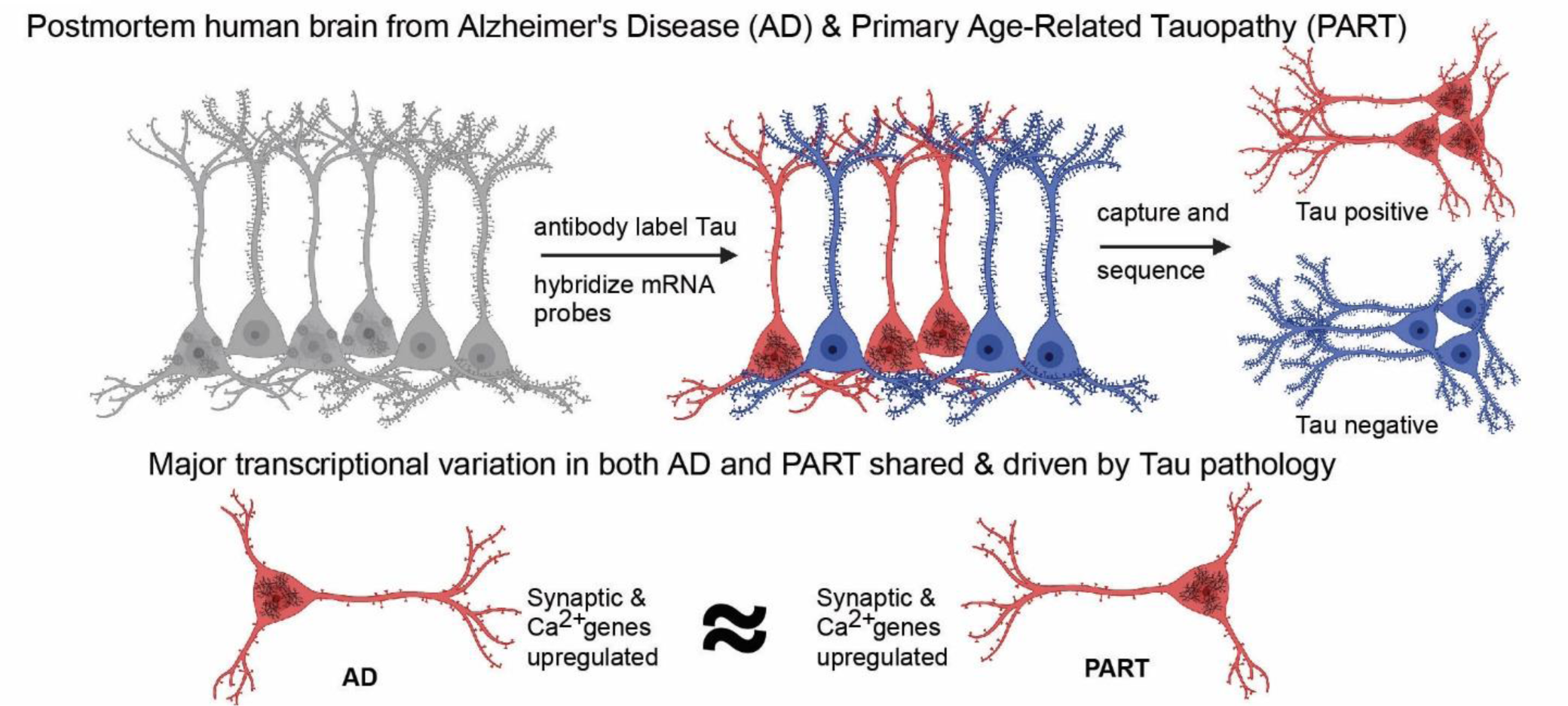

Created with BioRender.com (License GLSO).

## Background

Tau aggregates in neurons are present in the brain of nearly all elderly individuals[1, 2]. Many individuals show a characteristic accumulation of Alzheimer’s disease (AD) pathology with tau aggregates in neurons, called neurofibrillary tangles, and extracellular amyloid plaques, aggregates of the amyloid-beta peptide. Most remaining individuals show tau aggregates in neurons without significant amyloid plaques, called primary age-related tauopathy (PART)[3, 4]. However, little is known about the molecular changes associated with intraneuronal tau pathology in humans.

The relationship between the tau pathology in AD and PART is controversial. There are many similarities between the two conditions. The tau pathology in both diseases has a similar aggregate composition of hyperphosphorylated tau isoforms and an identical molecular structure of tau aggregates by electron microscopy[5]. Though tau pathology is common in human neurodegenerative diseases, no other sporadic neurodegenerative disease shows the same type of tau aggregate as is found in PART and AD. Additionally, the anatomic distribution and spread of tau pathology are also similar in the two diseases, leading to the use of the same tau pathologic staging system for both PART and AD[4, 6, 7]. Specifically, tau pathology accumulates early in the entorhinal cortex and hippocampus in both diseases, with later spread to the cortex. Cognitive impairment in both diseases is linked to the extent of tau pathology[4, 8-12]. These similarities have led some researchers to propose that PART may represent an early form of AD, prior to amyloid plaque formation[13].

However, the relationship between PART and AD is still unclear. Some studies have suggested that older individuals who are tau-positive and amyloid negative by imaging, possibly representing PART, often do not accumulate amyloid over time and have a different clinical progression from individuals who have imaging evidence of Alzheimer’s disease (tau-positive and amyloid positive)[14]. Genetic studies have highlighted distinct genetic risk factors for PART and AD[4, 15]. Furthermore, early PART cases have a higher predilection for the CA2 region of the hippocampus compared to AD, while AD cases tend to have higher levels of tau pathology which spreads farther in the brain [4, 16-18]. While there is evidence of synaptic loss in the hippocampus in both PART and AD, the overall pattern of synaptic changes may be distinct[16, 19-21]. So, despite the similarities between AD and PART, it remains to be determined whether the two processes are mechanistically related.

Despite extensive knowledge about the tau aggregates themselves in PART and AD, little is known about the molecular environment associated with intraneuronal tau in both diseases. Determining the transcriptional changes in human tissue specifically associated with intraneuronal tau pathology is technically challenging. Tau pathology accumulates only in a subset of neurons in the brain. Traditional bulk or single cell RNA sequencing methods are not able to segregate neurons with tau pathology from those without, thereby obscuring transcriptional signals associated with intraneuronal tau pathology. Prior studies have attempted to address this question in AD using laser capture microdissection[22], but these methods can only identify changes in the select genes present on microarrays. A single recent study developed a method of single soma sequencing with sorting by intraneuronal tau pathology using a phosphorylated tau antibody (AT8)[23]. Using this strategy, they identified transcriptional changes associated with tau pathology in cortical neurons in AD, most notably altered regulation of synaptic genes associated with intraneuronal tau. However, assessment of PART cases using this method would be challenging. This method would require frozen tissue of the hippocampus and entorhinal cortex, regions which are relatively small compared to cortical regions and popular regions to study in aging, potentially limiting the tissue available for study. The hippocampus and entorhinal cortex are also often affected by other pathologic processes in older individuals, such as ischemic changes, TDP-43 aggregation, and synuclein pathology[3, 11], which can be asymmetrically distributed between hemispheres [24] and may confound transcriptional analysis.

To investigate transcriptional changes associated with intraneuronal tau pathology in PART and AD, we leveraged Nanostring GeoMx spatial transcriptomic technology. This technology provides several advantages in querying transcriptional changes associated with tau pathology in the hippocampus and allows unbiased gene expression quantification [25, 26]. Specifically, paraffin-embedded hippocampal tissue sections are more readily available from human brain compared to hippocampal frozen tissue, and paraffin-embedded sections can be easily assessed using standard methods to exclude other common hippocampal pathologies, such as TDP-43, synuclein, or ischemic pathology. The GeoMx system also allows sub-segmentation of the region of interest using antibody labeling, so mRNA can be quantified from neurons with tau aggregation and adjacent neurons without tau aggregation in the same region, while excluding glial and other non-neuronal cell types from analysis.

In this study, we quantified transcriptional changes associated with intraneuronal tau pathology in the hippocampal CA1 region in AD and PART, and compared them to control cases. The AT8 antibody was used to identify neurons with tau pathology, a common antibody for labeling pathologic tau in human tissue[7]. Using this method, we identified differentially regulated genes in neurons with intraneuronal tau pathology in PART and AD, with striking similarities in the neuronal transcriptional regulation in both conditions. Using supervised and unsupervised methods, we identified synaptic gene changes associated with intraneuronal tau pathology in PART and AD. We were also able to identify gene expression signatures specifically associated with intraneuronal tau pathology. These findings were then verified using published single soma data[23] from cortical neurons with tau pathology in AD.

## Methods

### Human Tissue

All human tissue used in this study was from the Johns Hopkins Brain Resource Center (Baltimore, MD) with a post-mortem interval of less than 24 hours (Table 1). As tau pathology is a nearly universal feature of aging and most individuals over the age of 50 have tau pathology in the mesial temporal lobe [1, 2], control cases were defined as individuals under the age of 50 years with no evidence of amyloid deposition and no phosphorylated tau in the hippocampus or entorhinal cortex (PHF1, gifted from Dr. Peter Davies). PART cases were selected to have hippocampal tau pathology (Braak stage II-IV) with no amyloid deposition (Thal 0, CERAD 0). AD cases were selected for AD pathology (Braak V-VI, CERAD frequent) and were age-matched to the PART cases. All pathologic staging was performed used standard methods[7, 27, 28]. PART and AD cases were manually reviewed to ensure a sufficient number of tau-positive and tau-negative neurons in the CA1 region for transcriptomic evaluation. Cases were excluded from all categories of this study if there was α-synuclein pathology, TDP-43 pathology, frontotemporal dementia, or acute or chronic ischemic pathology in CA1.

**Table 1.** Case Information.

| Diagnosis (n) | Age | Sex (M:F) | PMI (hours) |
| --- | --- | --- | --- |
| Control (4) | 37.5 (36-39) | 2:2 | 17 (12-22) |
| AD (6) | 85.3 (75-94) | 3:3 | 9.1 (3-17.5) |
| PART (6) | 88.2 (77-95) | 4:2 | 12.9 (9-19) |

AD, Alzheimer’s disease; F, female; M, male; PART, Primary age-related tauopathy; PMI, post-mortem interval

### Antibodies

The AT8 (pS202/T205) phospho-tau (MN1020, Thermo Fisher Scientific, Waltham, MA) was used to label tau pathology in this study. Prior to use, the AT8 antibody was concentrated using a centrifugal filter (UFC9010, MilliporeSigma, Burlington, MA) then covalently labeled with Alexa 594 using an antibody labeling kit (A20185, Thermo Fisher Scientific, Waltham, MA) according to the manufacturer’s instructions.

### GeoMx Data Collection

Formalin-fixed, paraffin-embedded tissue sections were cut at 5 μm thickness from the mid-hippocampus at the level of the lateral geniculate nucleus. Sections were sent to the UPMC Hillman Cancer Center Flow Cytometry Core, where slides were prepared using standard GeoMx protocols (Manual Slide Preparation, MAN-10150-01) for the Nanostring GeoMx whole transcriptome atlas (HS_R_NGS_WTA_v1.0). The labeled AT8 was used at a dilution of 1:800 as a morphology marker for tau pathology. Slides were loaded into the GeoMx DSP instrument and scanned using default instrument settings. Experimenters were blinded to disease group. 2-7 regions of interest (ROI) (660 × 785 μm) were selected within the CA1 region of the hippocampus per case. Regions were selected to include a minimum of 5 neurons per group (control: no tau, disease: with and without tau) while avoiding areas artifactual tissue disruption. The ROI image was exported to Image J, a macro was run to set up the masking of each selected neuronal group (Add. File 1), and neurons for each group were manually selected. As only cell bodies could be visualized using this method, only neuronal cell bodies were selected for collection. Pyramidal excitatory neuron cell bodies were identified in the CA1 pyramidal cell layer by morphological features, including large triangular cell body shape, large nucleus, and a prominent nucleolus, and they were segmented by whether the cell body contained AT8-positive tau pathology. Preliminary experiments with a chemical lipofuscin quenching agent (TrueBlack Plus, Biotium, Fremont, CA) showed reduced mRNA counts in treated sections (data not shown), so no lipofuscin quench was used in quantified experiments. A subset of neurons were excluded due to an indeterminate fluorescent signal which was not definitely lipofuscin or low level phosphorylated tau. The manual selection for each group was exported as a PNG file, and uploaded into the GeoMx ROI as a mask. Selections were collected iteratively by the GeoMx from the ROIs into a 96 well plate. Collected products were processed and sequenced at the University of Pittsburgh Health Sciences Sequencing Core. Requested sequencing depth was calculated from the total collected area. Collected product was sequenced on an Illumina NextSeq 2000 (Illumina, Inc., San Diego, CA). Resulting FASTQ files were decoded and processed into count files using the NanoString GeoMx NGS Pipeline app (version 2.0.21) in the Illumina BaseSpace Sequencing Hub. Count files were uploaded back to the GeoMx DSP instrument indexed with the corresponding slide scans for analysis.

### Collection and Quality Control

Collected cells per segment were quantified manually. For each dataset, segment properties were exported from the GeoMx and the deduplicated reads were divided by the cell count for that segment to get the deduplicated reads per cell (Fig. S1). mRNA quality and gene expression analysis were performed in R statistical software (version 4.2.2)[29] / RStudio (Posit, Boston, MA). Quality analysis was performed using the Nanostring GeoMx packages on Bioconductor (NanoStringNCTools version 1.4.0, GeomxTools version 3.0.1, GeoMxWorkflows version 1.2.0)[30-32]. All segments passed quality control metrics for percent trimmed reads, stitched reads, aligned reads, and sequencing saturation (>85% all measures). Segments were excluded from analysis for low gene detection rate (<2% of whole transcriptome atlas), which included 2 AD tau-negative segments and 9 AD tau-positive segments spread across 3 separate AD cases. Genes were included in the quantitative analysis if they were greater than 2 standard deviations above the GeoMean of the negative control probes in more than 20% of segments. Normalization was done using the calcNormFactors function from the edgeR Bioconductor package as previously described [33-35]. Briefly, the trimmed mean of M-values (TMM) between each pair of samples was calculated to account for differences in library sizes between samples. As day of collection remained the greatest source of variation in the data, ComBat batch correction was applied using the three separate profiling days as the batch variable[36, 37]. Principal components analysis and UMAP dimensionality reduction was performed to visualize the corrected data with regard to slide, sex, and collection (Fig. S2)[38]. While slide, disease, and tau pathology remained significant sources of variation, sex was not. Additional Combat correction was performed for both collection day and slide on data to be used for downstream dimension reduction analysis, i.e. PCA and CoGAPS analysis[39]. However, because of concerns over confounding and the ability to account for slide effects, the data with only day of collection Combat batch correction was used for differential expression analysis.

### Differential Gene Expression Analysis

Differential gene expression was performed using the lme4 Bioconductor packages in the R programing language as previously described [40]. Briefly, two linear mixed-effect model (LMM) were fit to the data. In both models, slide was included as a random intercept term to account for any technical variation. The first model tested for the presence of tau-negative versus tau-positive neurons in the context of both AD and PART. The second model tested for genes that were differentially expressed between AD, PART, and control. Differentially expressed genes in all analyses were defined as a log2 fold change greater than 0.2 and a false discovery rate-corrected *p* value of less than 0.05. Initial gene functional annotations were based on GeneCards [41] and ShinyGO (version 0.77) [42] annotations.

### Synaptic (SynGO) Analysis for Gene Expression Changes

The list of quantified genes for hippocampal neurons and the literature cortical single cell dataset were entered into the synaptic gene ontology database (SynGO, https://www.syngoportal.org/)[43] and genes with annotations in SynGO were used to define the class of synaptic genes. The distribution of the fold change for each comparison was plotted in GraphPad Prism 9.4.1 (GraphPad Software, San Diego, CA) and fit using a Gaussian model with least squares regression. The curve fit of synaptic genes were either compared to a theoretical mean of 0 (no change in that comparison) or to the mean of the fit of the distribution of all quantified genes for that dataset using an extra sum of squares F test. Comparison with a theoretical mean of 0 was used in the hippocampal dataset because the total quantified gene changes was not different from 0 for any comparison. Comparison to the overall quantified gene distribution was used for the cortical single cell dataset as this was significantly different from 0 in some cell types.

### CoGAPS and PatternMarkers Analysis

Coordinated Gene Activity in Pattern Sets (CoGAPS) analysis was applied to the entire log transformed Combat corrected gene expression dataset and run for a range of dimensionalities. Stability of the patterns and the composition of the associated gene expression signatures was assessed as previously described [44] and a dimensionality of five was chosen. Patterns were visualized to assess the ability to distinguish between Tau status, sex, collection date, slide, and disease status. *Post hoc* statistics were used to quantify the significance of observed differences in sample weights. The SynGO gene list generated for DE was use as input to Fast Gene Set Enrichment Analysis (fsea) for enrichment analysis of the gene loadings for all five patterns [45]. The PatternMarker statistic was calculated on the corresponding gene loadings of all five patterns generating lists for use as data driven biomarkers for each pattern [39]. Gene ontology analysis was conducted using ShinyGO (version 0.77)[42].

### Projection Analysis

Single soma data[23] from cortical neurons with tau pathology in AD was projected into all five CoGAPS signatures learned on the GeoMx data (described above) using the projectR Bioconductor package [44, 46]. Briefly, the log transformed preprocessed expression data was stratified by excitatory and inhibitory neuron types, subset to those genes common to both datasets, and input to the projectR function along with the feature loadings from all five CoGAPS patterns. The resulting projected pattern weights for the single soma data were then visualized by each cell type, disease state, and Tau status using the annotation reported in [23]. *Post hoc* statistics assessing the differential distribution of projected pattern weighs across cell type, disease state, and Tau status were used to quantify the significance of observed differences in the use of each CoGAPS signature and the biological processes associated with it.

### Statistical Analysis

Gaussian modeling with least squares regression, simple linear regressions, and extra sum of squares F tests were performed using GraphPad Prism 9.4.1 (GraphPad Software, San Diego, CA). Linear mixed model regression, PCA, CoGAPS analysis, projectR analysis, fgsea, t-tests, and anova analysis were performed in the R statistical language version 4.2.2 [47].

## Results

### mRNA Quantification in Neurons with Intraneuronal Tau Pathology

Information about transcriptional regulation in neurons with intraneuronal tau pathology could provide key insights into pathogenic mechanisms associated with tau aggregation in PART and AD, and into the relationship between tau pathology in the two conditions. However, little is known about the transcriptional changes associated with intraneuronal tau pathology in Alzheimer’s disease (AD). The transcriptional changes in the related primary age-related tauopathy (PART) are essentially unknown. To investigate transcriptional changes associated with intraneuronal tau pathology in AD and PART, we used GeoMx digital spatial profiling of hippocampal CA1 neurons with intraneuronal tau pathology or lacking intraneuronal tau pathology. Unlike other transcriptomic methods, the GeoMx method allows for collection of mRNA information from selected cells within a region of interest enabling quantification of mRNA expression from subsets of neurons within an anatomic region (Fig. 1A).

**Figure 1.**
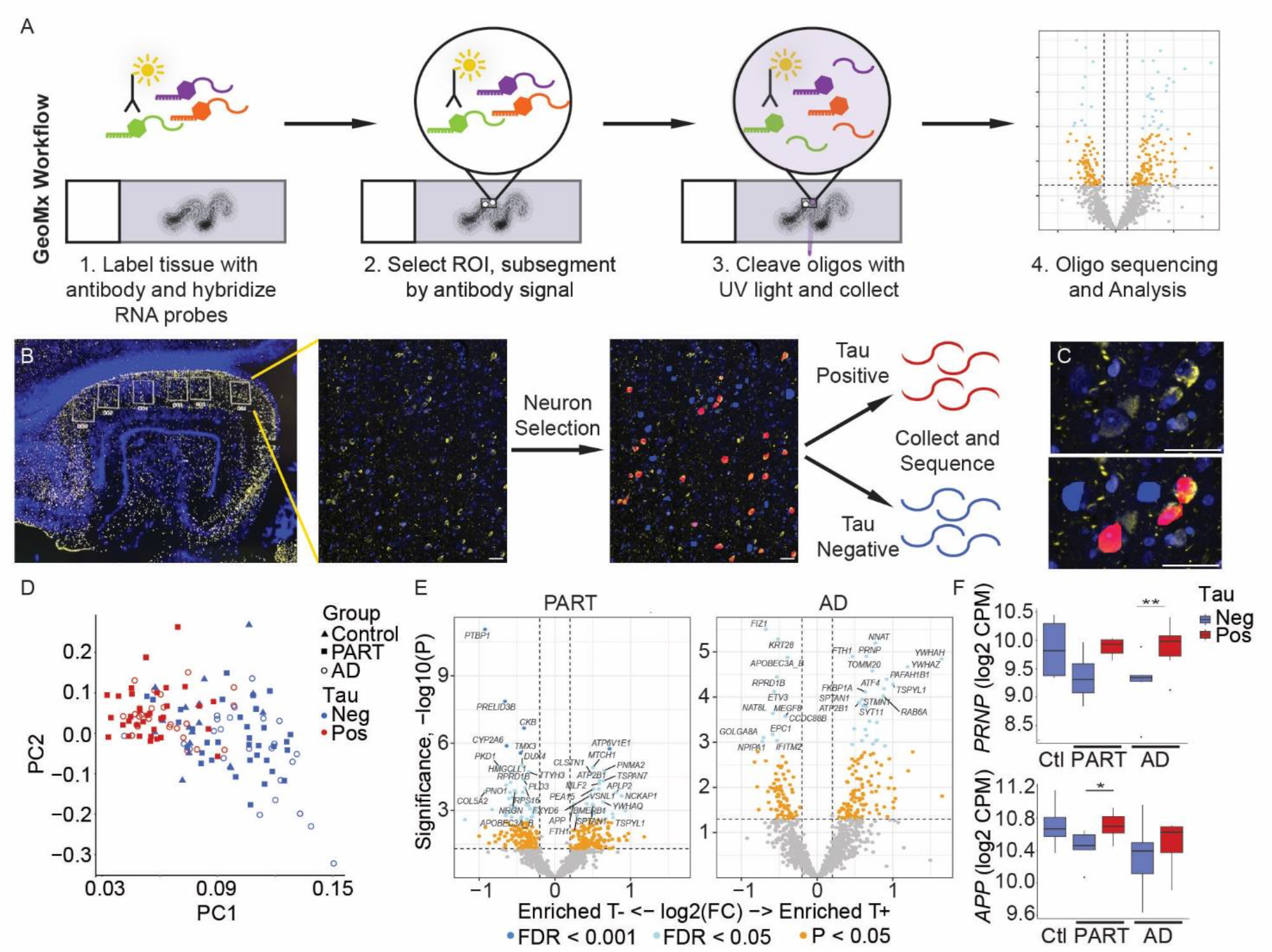
Transcriptional Changes Associated with Intraneuronal Tau Pathology in PART and AD. (A) GeoMxWorkflow. First, tissue sections are labeled with antibodies and hybridized with GeoMx mRNA probes. These probes are linked with a unique oligonucleotide barcode via a chemical linker which can be cleaved using ultraviolet light. A region of interest (ROI) is selected on the tissue, then segments are selected within the region of interest by antibody labeling. A 10μm ultraviolet light is used to cleave the barcodes within each selected segment, which are then collected, sequenced, and analyzed. Barcode collection can be done iteratively within an ROI, so multiple segments with differential antibody labeling can be collected from a single ROI. (B) In this study, hippocampal sections were stained for phosphorylated tau to mark tau pathology (yellow, AT8) and counterstained with a marker for nucleic acid (blue, SYTO13). Regions of interest were selected within the CA1 region on the Nanostring GeoMx, then regions were segmented and masked based on neurons with tau pathology (red, tau-positive) or without (blue, tau-negative). Transcript-specific oligonucleotide barcodes were then collected and sequenced from each selected segment. (C) High power view of neurons selected as positive (red) and negative (blue) for tau pathology. (D) Principal component analysis of each segment by disease group and tau status. (E) Differentially expressed genes in tau-positive versus tau-negative neurons in PART and AD. (F) Expression of representative genes previously reported as altered in tau-positive neurons. Significance based on mixed effects linear modeling of differentially expressed genes after false discovery rate correction. AD, Alzheimer’s disease; Ctl, control; CPM, counts per million; FC, fold change; FDR, false discovery rate; Neg, tau pathology negative; Oligo, oligonucleotide; PART, primary age-related tauopathy; PC, principal component; Pos, tau pathology positive; T-, tau pathology negative; T+, tau pathology positive. Scale bars are 50 μm. *FDR<0.05, **FDR<0.01.

In this study, hippocampal CA1 pyramidal neurons with and without intraneuronal tau pathology were collected from 6 AD cases, 6 PART cases, and 4 control cases (Table 1). The CA1 region was chosen due to the high burden of tau pathology in this region in PART and AD, and the relatively uniform composition of excitatory pyramidal neurons in the CA1 pyramidal layer within a transverse section[48]. Pyramidal excitatory neurons were segmented as positive or negative for intraneuronal tau pathology, which was highlighted by phospho-tau immunofluorescence using the AT8 antibody (Fig. 1B,C), an antibody commonly used for diagnostic evaluation of tau pathology in PART and AD [4, 7]. For disease cases an average of 12-21 neurons were collected per segment, defined as one group of neurons within a region of interest with the same tau status (Fig. S1A). As there was only one neuronal population collected in control cases (tau-negative), there were on average twice as many neurons collected in control segments. Despite the low number of neurons collected per segment, good quality mRNA quantification was obtained (Fig. S1B,C). Average quantified mRNA was similar between groups regardless of disease or tau status (Fig. S1D), with an average of 6,310 mRNA counts per cell after de-duplication of redundant oligonucleotide barcodes. However, there was greater variability in the quantified mRNA per cell within the control group compared to disease groups. mRNA counts did not vary by the time tissue spent in a paraffin block, up to nearly 17 years, nor did it vary by post-mortem interval, up to 22 hours (Fig. S1E,F).

Concordant with the relatively few neurons collected in each segment, approximately 2-10% of the whole transcriptome could be quantified in most segments (Fig. S1G). Segments with less than 2% quantifiable gene expression were excluded from further analysis. After curating genes for sufficient quantifiable expression, 1392 genes were analyzed. Quantified genes were enriched in genes expressed by excitatory neurons, but not enriched for genes expressed in astrocytes, oligodendroglia, or microglial/myeloid lineage cells (Fig. S3). This gene set was also enriched for genes with synaptic annotations (353 genes annotated in SynGO, 25.4% of quantified genes), consistent with collection of mRNA specifically from neurons.

After batch correction, the largest variation in gene expression in this dataset was associated with intraneuronal tau pathology by principal component analysis (Fig. 1D). Tau-positive neurons separated from tau-negative neurons in both PART and AD, with control neurons largely centered between the two groups. Notably, segments from PART and AD did not separate by disease group in principal components 1 and 2. Therefore, in hippocampal neurons at least, gene expression variation was more associated with intraneuronal tau pathology than with disease (PART or AD).

### Gene Expression Changes Associated with Intraneuronal Tau in PART and AD

Multiple genes showed differential expression in tau-positive neurons compared to tau-negative neurons in PART and AD (Fig. 1E). Mixed effects linear modeling was used to determine differential gene expression, with differentially expressed genes (DEGs) defined as having a log2 fold change greater than 0.2 and a false discovery rate-corrected p value of less than 0.05. In PART, 72 genes were differentially expressed in tau-positive neurons, with 26 genes upregulated and 46 genes downregulated (Add. File 2). In AD, 39 genes were differentially expressed in tau-positive neurons, with 27 genes upregulated and 12 downregulated (Add. File 2). This higher proportion of upregulated DEGs in tau-positive hippocampal neurons in AD (69%) is similar to what has been reported previously in cortical neurons [23]. Several DEGs in tau-positive neurons in either PART or AD had been reported previously as DEGs in tau-positive neurons in the cortex in AD [23]. This included the prion protein gene (*PRNP*) and amyloid precursor protein gene (*APP*), critical genes in human neurodegenerative disease, which were upregulated in tau-positive neurons in AD and PART, respectively (Fig. 1F).

Analysis of the DEGs in tau-positive neurons in PART and AD showed several interesting trends. Tau-positive neurons in PART showed differential expression of genes in the amyloid precursor protein (*APP*) family and genes involved in APP processing (*APP, APLP2, PLD3, CLSTN1*) [49-51], as well as genes involved in calcium regulation and signaling (*TTYH3, VSNL1, ATP2B1, CALM1*) and mitochondrial function (*TXN2, TOMM5, SARDH1, VDAC3, MTCH1*). Multiple genes involved in mRNA regulation, transcription, and splicing were down-regulated in tau-positive neurons in PART (*PTBP1, PNO1, RPRD1B, PNN, DUX4*). Tau-positive neurons in AD showed up-regulation of several pre-synaptic genes (*VAMP2, SYT1*) and 14-3-3 family members (*YWHAG, YWHAH, YWHAZ*), which have previously been implicated in AD and associated with tau pathology in AD [52]. Multiple genes implicated in apoptosis, ER stress, and stress granule pathways were differentially regulated in PART and AD (PART: *MTCH1, PEA15, PNMA2, SPIN2B, TXN2, SCAMP5*; AD: *IFITM2, CCDC88B, PTMA, G3BP2, ATF4*) [41].

DEG analysis comparing disease to control samples was limited by the small sample numbers and high variability in the control samples. Only 20 genes were identified as DEGs in AD and only 9 DEGs were identified in PART compared to controls (Add. File 3). Several DEGs (9/20) in AD were previously reported in excitatory neurons in a variety of brain regions [53, 54], 7 of which showed the same directional change in published datasets (*LMO4, PPP2CA, GOT1, ATP2Ba, RAB3A* down-regulated; *CXCR4, PXN* up-regulated). Of the 9 DEGs identified in PART, 8 were also present in the AD-associated DEGs with the same directional change, suggesting these could be changes associated with aging in both conditions. One DEG, *JOSD2*, was up-regulated in excitatory neurons in PART. This gene showed a similar trend toward up-regulation in AD which did not quite reach significance (*p*=0.07 after FDR correction). Therefore, while somewhat underpowered, this analysis did not identify any DEGs which appeared to be specific to PART.

### Synaptic Gene Regulation Associated with Intraneuronal Tau Pathology

Synaptic genes have been shown to be dysregulated in AD specifically in neurons with intraneuronal tau pathology in the cortex [23, 54], so we examined whether synaptic genes were similarly dysregulated in PART and AD in the hippocampus. Given the relatively small population of genes sampled in this dataset, conventional gene ontology analysis was of limited utility. Instead, the intersection of quantified genes and those in the SynGO database[43] was used to identify 353 genes with synaptic annotations. The fold change of these synaptic genes was analyzed either by comparing control cases to PART and AD (Fig. 2A-C, Add. File 3, 4), or by comparing tau-positive to tau-negative neurons within PART or AD (Fig. 2D-F, Add. File 3, 4). When all neurons within a disease group were combined (tau-positive and tau-negative), synaptic genes were downregulated in PART neurons compared to control neurons (Fig. 2B, mean log2 fold change -0.12). A greater down-regulation of synaptic genes was present in AD neurons compared to control neurons (Fig. 2C, mean log2 fold change - 0.18). This downregulation is similar to prior reports showing synaptic gene downregulation with aging and AD in multiple brain regions, including hippocampus [54]. However, when synaptic gene expression was compared in tau-positive versus tau-negative neurons, the findings were different. Synaptic genes were up-regulated in tau-positive neurons in PART (Fig. 2E) and in AD (Fig. 2F). This effect again showed a greater magnitude in AD (mean log2 fold change 0.13) compared to PART (mean log2 fold change 0.073). Analysis of all the quantified genes showed no evidence of global transcriptional changes by disease state or by intraneuronal tau pathology (Fig. S4A-D). Examination of individual gene expression data of representative synaptic (SYT1) and calcium signaling (CALM3) genes show down-regulation of these genes in disease tau-negative neurons, with relative preservation of gene expression in disease tau-positive neurons (Fig. 2G,H).

**Figure 2.**
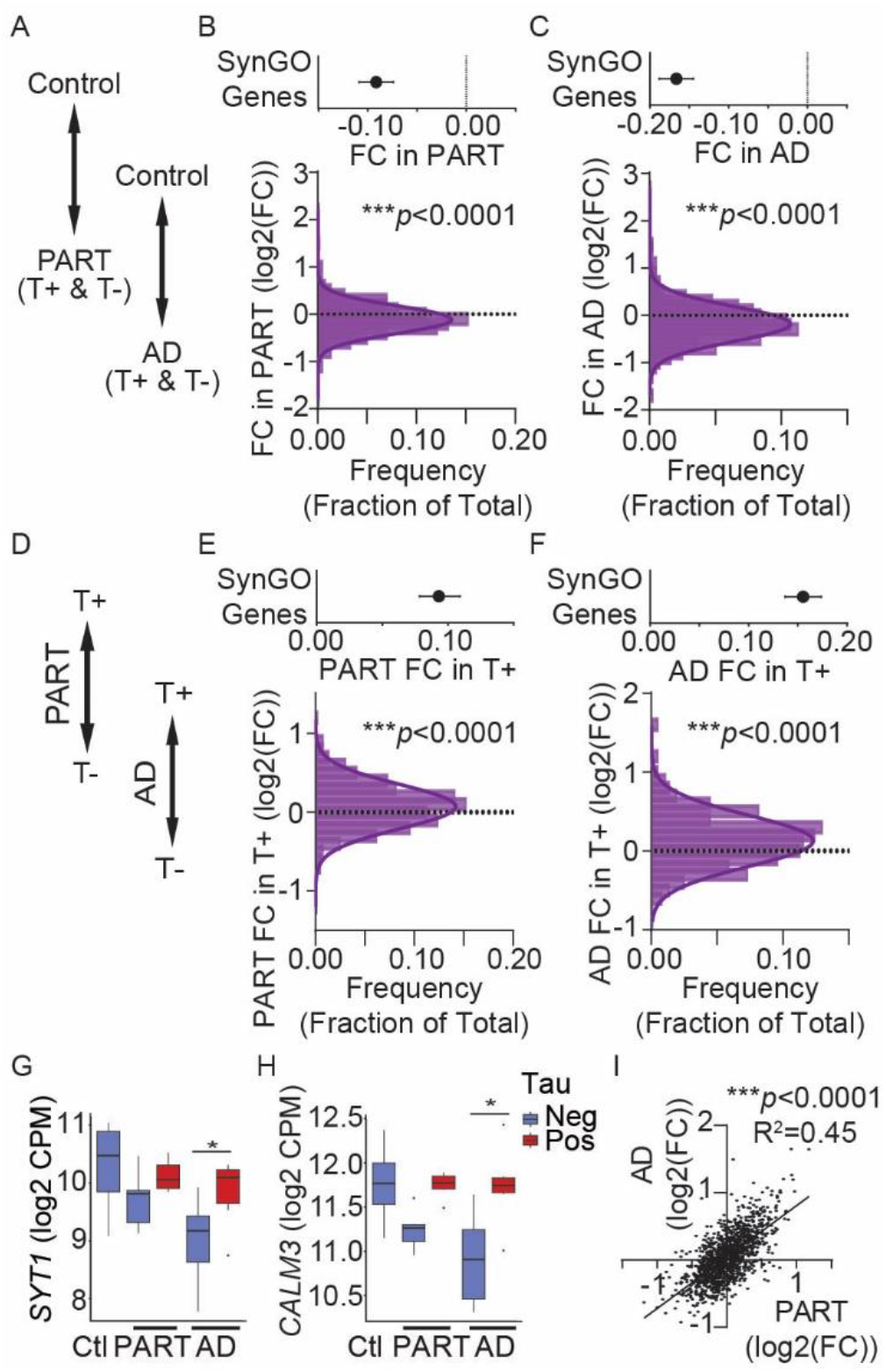
Synaptic Gene Changes Associated with Intraneuronal Tau Pathology in PART and AD. All quantified genes were processed in SynGO to identify genes with synaptic annotations. (A) Schematic of group comparison of synaptic gene expression changes in B and C; the control group was compared either to all neurons in PART or to all neurons in AD to mimic effects seen in traditional bulk analysis. (B) Decreased synaptic gene expression in PART versus control shown by mean fold change of SynGO genes (top) with histogram (below). (C) Decreased synaptic gene expression in AD versus control shown by mean fold change of SynGO genes (top) with histogram (below). (below). (D) Schematic of comparison of synaptic gene expression changes in E and F. Tau-positive neurons were compared to tau-negative neurons in PART or AD. (E) Increased synaptic gene expression in tau-positive neurons in PART (E) and AD (F) shown by mean fold change of SynGO genes (top) with histogram (below). (G,H) Representative gene expression changes by disease group and tau status. Significance by mixed effects linear model of differential gene expression after false discovery rate (FDR) correction. *FDR<0.05. (I) Log2 of the fold change in tau positive versus negative for each quantified gene in PART (x-axis) and AD (y-axis) showing a significant correlation (simple linear regression, *p*<0.0001, R^2^=0.45). AD, Alzheimer’s disease; Ctl, control; CPM, counts per million; FC, fold change; Neg, tau pathology negative; PART, primary age-related tauopathy; Pos, tau pathology positive; T-, tau pathology negative; T+, tau pathology positive. B,C,E,F – curve generated by Gaussian least squares fit and significance is mean significantly difference from 0 by extra sum of squares F test; dotted line at 0. Error bars are SEM. G,H – Error bars are 95% CI.

As similar trends were present in tau-positive neurons in PART and AD, we analyzed whether overall gene expression changes associated with intraneuronal tau pathology were similar between the two diseases. The log2 fold change for all quantified genes in tau-positive neurons was plotted for PART (x-axis) and AD (y-axis) (Fig. 2I). There was a striking correlation between the fold change associated with intraneuronal tau pathology across all 1392 quantified genes (simple linear regression, *p*<0.0001, R^2^=0.45), implying that intraneuronal tau pathology is associated with similar transcriptional regulation in PART and AD.

For an unsupervised analysis of gene expression patterns, coordinated gene association in pattern sets (CoGAPS) analysis was performed followed by PatternMarkers analysis to identify the associated genes in each pattern. CoGAPS analysis was used to generate 5 coordinated gene expression patterns (Add. File 5). Two of these gene expression patterns were associated with intraneuronal tau pathology. The first pattern showed significant up-regulation associated with intraneuronal tau (tau+/AD vs tau-/AD: t_(46)_ = 10.52, *p* = 7.94e-14; tau+/PART vs tau-/PART: t_(67)_ = 8.4518, *p* = 3.69e-12) (Fig. 3A,B), with similar changes in PART and AD (tau+/AD vs tau+/PART: t_(56)_ = -0.021, *p* = 0.983; tau-/AD vs tau-/PART: t_(57)_ = -0.532, *p* = 0.5967). There were 164 patternMarker genes for with the tau up-regulated pattern. This pattern was notable for genes involved in neuronal calcium signaling (*CALM1, CALM2, CALM3, CAMKK1*, CAMK2B) and was enriched for SynGO-annotated genes (Fig. S5, *p*=.0001), similar to what was seen in the supervised analysis. GO analysis of Biological Processes using ShinyGO[42] showed enrichment in this up-regulated pattern for synaptic signaling pathways (FDR=1.4×10^-4^) and pathways involved in calcium ion transport and cation homeostasis (FDR=1.4×10^-4^) (Fig. 3C, Add. File 6). The highest enrichment was seen in regulation of synaptic vesicle exocytosis pathways (5-fold enrichment, FDR=1.5×10^-4^), implying a regulation of pre-synaptic function by intraneuronal tau pathology.

**Figure 3.**
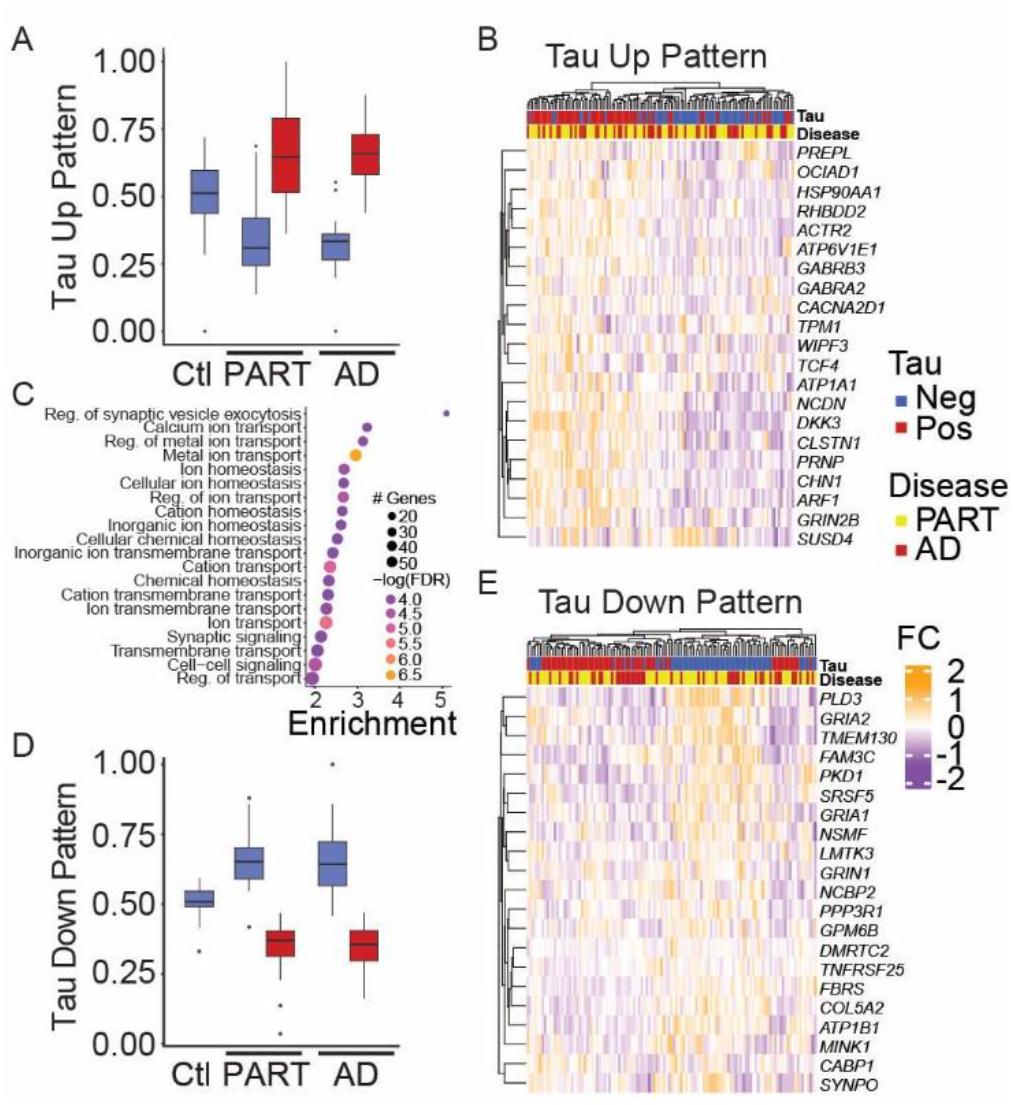
Coordinated Gene Association in Pattern Set (CoGAPs) Analysis by Intraneuronal Tau Pathology. CoGAPS analysis was performed on the quantified genes with 2 patterns showing variation by intraneuronal tau pathology. (A) Coordinated gene expression score for Tau Up pattern by disease group and tau pathology and (B) expression heatmap of the top 20 genes in this pattern by tau and disease group selected by score after PatternMarkers analysis. (C) The genes significantly associated with the Tau Up pattern after PatternMarkers analysis were analyzed for enriched GO Biological Processes using ShinyGO [42] and the results are displayed as a dot plot. The size of each dot reflects the number of pattern genes in the process, while the color reflects the *p* value after false discovery rate correction. (D) Coordinated gene expression score for Tau Down pattern by disease group and tau pathology, and (E) expression heatmap of the top 20 genes in this pattern by tau and disease group selected by score after PatternMarkers analysis. AD, Alzheimer’s disease; Ctl, control; FC, log2 fold change; FDR, false discovery rate corrected *p* value; Neg, tau pathology negative; PART, primary age-related tauopathy; Pos, tau pathology positive; Reg., regulation. B,D - Error bars are 95% CI.

The second coordinated gene expression pattern was significantly down-regulated in tau-positive neurons (tau+/AD vs tau-/AD: t_(46)_ = -10.07, *p* = 3.21e-13; tau+/PART vs tau-/PART: t_(67)_ = - 13.91, *p* < 2.2e-16) (Fig. 3D,E) with similar changes in PART and AD (tau+/AD vs tau+/PART: t_(56)_ = 0.057, *p* = 0.955; tau-/AD vs tau-/PART: t_(57)_ = -0.011, *p* = 0.991).

This tau down-regulated pattern has 127 patternMarker genes, though the biologic significance of this pattern was less clear. Gene ontology analysis was showed no significant pathway enrichment in this gene set. Closer examination of the function of these genes showed a mix of genes involved in synaptic function, mitochondrial function, metabolism, and mRNA regulation. While many synaptic genes were in the up-regulated pattern, several important synaptic genes were associated with the down-regulated pattern, including glutamate receptors (*GRIA1, GRIA2, GRIN1*) and several genes involved in synaptic signaling pathways (*PRKCA, CAMK2A, CACNG8, DLG2/PSD-93, NRXN2*). The other three coordinated gene expression patterns showed no clear association with either intraneuronal tau or disease (Fig. S4E-G).

### Shared Signatures of Intraneuronal Tau in Cortical Excitatory Neurons

To determine if these gene expression changes were reproducible across independent datasets and thus generally associated with intraneuronal tau in AD, we compared our findings to a single soma RNA-seq dataset from cortical neurons in AD sorted by intraneuronal tau pathology [23]. First, changes in synaptic gene expression were assessed. Genes quantified in the analysis of intraneuronal tau pathology in the published dataset were analyzed for SynGO[43] annotation for each cell type. To analyze only excitatory neurons with a significant proportion of tau pathology, neurons were grouped as they were analyzed in the original publication[23]. For each excitatory neuron group, the fold change of synaptic genes in tau-positive neurons was compared to the fold change of all measured genes in tau positive neurons. Of 9 excitatory neuronal subtypes with intraneuronal tau pathology, 7 groups of excitatory neurons showed significant upregulation of synaptic genes in tau-positive neurons (Fig. 4A, S6) while two excitatory subtypes showed no change. No excitatory neurons showed down-regulation synaptic genes in tau-positive neurons.

**Figure 4.**
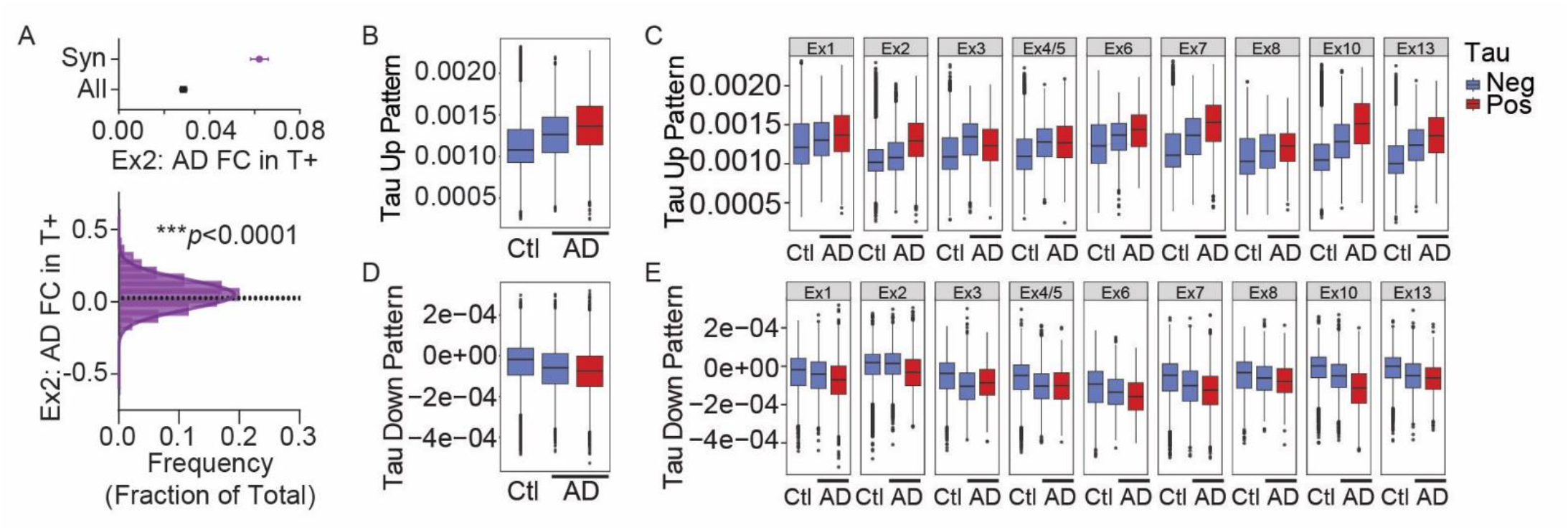
Synaptic and Gene Expression Patterns Associated with Cortical Intraneuronal Tau in Single Cell Dataset. Publicly available single soma RNA-seq data from cortical neurons in AD [23] was analyzed for synaptic gene expression changes and expression of the Tau Up and Tau Down patterns. Excitatory neuronal grouping was performed as in the published analysis for tau positive and negative neurons. (A) Representative plots of increased synaptic gene expression (SynGO annotated) compared to expression of all quantified genes in excitatory neuron group 2 (Ex2: *CUX2-COL5A2*) shown by mean fold change of genes with SynGO annotations (top) with associated histogram (below). Curve generated by Gaussian least squares fit and significance is means of synaptic genes and all measured genes are different by extra sum of squares F test; dotted line at mean of all measured genes in Ex2. (B) Increased expression of Tau Up pattern in overall tau-positive excitatory cortical neurons and (C) by excitatory neuronal group. (D) Decreased expression of Tau Down pattern in overall tau-positive excitatory cortical neurons and (E) by excitatory neuronal group. AD, Alzheimer’s disease; All, all quantified genes; Ctl, control; FC, log2 fold change; Neg, tau pathology negative; Pos, tau pathology positive; Syn, synaptic genes; T+, tau-positive. A – Error bars are SEM. B-E - Error bars are 95% CI.

Next, the published AD cortical neuron dataset was projected into the identified coordinated gene expression patterns identified in PART and AD using the transfer learning algorithm ProjectR. In the single soma cortical AD data, the gene expression pattern upregulated in tau-positive hippocampal neurons (Tau Up) was also significantly upregulated in tau-positive cortical neurons in AD (t_(91949)_ = 70.419, *p* < 2.2e-16) (Fig. 4B). When excitatory cortical neurons were separated by group (Fig. 4C), this effect was most prominent in in 4 of the 5 groups of cortical neurons with highest levels of tau pathology (increased in Ex1, Ex2, Ex7, Ex10, but not in Ex3)[23]. When the down-regulated pattern in tau-positive hippocampal neurons (Tau Down) was examined, similar effects were identified both globally and on with cell type specific interactions. This gene expression pattern was significantly downregulated in tau-positive cortical neurons in AD overall (t_(91949)_ = -36.641, *p* < 2.2e-16), and the effects were most prominent in excitatory neuronal groups with the highest proportion of tau pathology (Fig. 4D,E).

## Discussion

Little is known about the gene expression changes associated with intraneuronal tau pathology in the hippocampus in human Alzheimer’s disease (AD), and even less is known about gene expression changes associated with tau pathology in primary age-related tauopathy (PART). Here we used spatial transcriptomic technology to identify hippocampal gene expression changes associated with intraneuronal tau pathology in AD and PART. Tau pathology was the major factor associated with hippocampal neuronal gene expression changes in PART and AD. Surprisingly, gene expression changes were similar in tau-positive neurons in PART and AD. When examining synaptic gene expression, synaptic genes were downregulated in PART and AD overall, but tau-positive neurons in disease showed relatively higher synaptic gene expression compared to tau-negative neurons in disease. Unbiased coordinated gene expression in pattern sets (CoGAPS) and PatternMarkers analysis identified two gene expression patterns associated with intraneuronal tau in AD and PART. The gene expression pattern up-regulated in tau-positive neurons was enriched for synaptic genes, specifically genes involved in pre-synaptic vesicle exocytosis, and genes in calcium regulation pathways. Finally, both the synaptic gene expression changes and the tau-associated gene expression pattern findings were replicated in published independent data of tau-positive cortical neurons in AD.

To our knowledge, this is the first study of tau-associated gene expression changes in PART. In older individuals without AD pathology, PART is a nearly universal finding at autopsy[3, 4]. In some cases with higher tau pathology [4, 9, 10, 12, 55], PART is associated with cognitive impairment. However, studies of gene expression changes in PART are lacking. Here we show robust gene expression changes in PART associated with intraneuronal tau pathology. Overall, PART is associated with decreased synaptic gene expression compared to controls, similar to what has been described previously in the hippocampus with aging [54]. However, neurons with tau pathology in PART showed increased synaptic gene expression and alterations in genes associated with calcium regulation and signaling. Intraneuronal tau in PART was also associated with altered regulation of *APP, APLP2* (a related gene in the APP family), and altered expression of several genes involved in APP processing (*PLD3, CLSTN1*). This pathway is critical to the cleavage of *APP* into the amyloid beta peptide which accumulates into extracellular amyloid plaques in AD[49-51]. While no prior studies are available for PART, prior studies in AD have also shown increased *APP* expression in tau-positive neurons at the mRNA and protein level[23, 56]. While the result of these gene expression changes on amyloid production in PART is not clear, these APP pathway changes suggest a link between intraneuronal pathology and altered processing of *APP*.

The gene expression changes associated with intraneuronal tau in PART and AD are strikingly similar. There is controversy over whether PART and AD are related[3, 13]. Despite similarities in tau aggregates and anatomic progression of the two diseases[4-7], there are significant genetic and pathologic differences[4, 15, 17, 18, 57] and the relationship between the tau pathology in the two conditions is unclear. The findings in this study show clear transcriptional similarities associated with intraneuronal tau pathology in PART and AD, implying that there may be a mechanistic link between the tau pathology in the two conditions. However, similar transcriptional studies have not been performed in other types of tauopathies. Thus, it remains to be determined whether this transcriptional signature reflects tau aggregation in neurons generally or whether it is specific to the type of tau aggregation found in PART and AD.

The synaptic gene expression findings in this study suggest a re-evaluation of how we think about synaptic gene regulation in PART and AD. The decreased synaptic gene expression comparing disease to control cases is entirely expected and replicates prior studies on gene expression in aging and AD[54, 58]. However, this decrease in synaptic gene expression was driven primarily by neurons lacking intraneuronal tau pathology. Tau-positive neurons showed increased synaptic gene expression compared to tau-negative neurons in disease. These synaptic gene changes were confirmed in the single comparable study of tau-positive cortical neurons in AD. Interestingly, while very few studies in preclinical models examine gene changes associated with intraneuronal tau pathology, rare studies in tau transgenic mice have shown increased expression of select synaptic genes[59, 60].

Unsupervised analysis of gene expression patterns confirmed that synaptic genes, particularly those involved in synaptic exocytosis, and genes involved in calcium signaling and regulation were upregulated in tau-positive neurons in PART and AD. This gene expression pattern was confirmed in excitatory cortical neurons in AD, despite a different methodology and different brain region assessed in the independent comparison dataset. This regulation of synaptic gene expression therefore appears to be a generalized response in most excitatory neuron types with high levels of tau pathology. However, the significance of this synaptic gene upregulation remains unclear. Given that individual cortical neurons with tau pathology in AD have been shown to lose synaptic spines[61] and that high levels of tau pathology are associated with evidence of synaptic loss in PART and AD[16, 19, 56, 62, 63], the synaptic gene upregulation may represent a neuronal compensatory response to synaptic loss. It is also possible that these synaptic gene changes in tau-positive neurons are a compensatory response which may facilitate neuronal survival, as prior studies have shown that the neuronal populations with a high tau pathology burden are not the most susceptible to cell death in AD and that cortical neuronal loss far exceeds the population of neurons with tau pathology[23, 64].

The reason for the down-regulation of synaptic genes in PART and AD in tau-negative neurons is profoundly unclear. Prior literature has shown downregulation of synaptic genes in human brain with aging in multiple brain regions, which is exacerbated by AD[54], consistent with the findings in this study. In both AD and PART, cortical regions only show a smaller subset of neurons affected by tau pathology[23, 65]. Therefore, prior studies showing a down-regulation of synaptic genes in cortical transcriptional data would largely represent tau-negative neurons. Interestingly, the age-associated down-regulation of synaptic and neuronal genes is reported to be similar in humans and rhesus macaques [66] and synaptic gene down-regulation via the REST transcription factor pathway has been associated with increased longevity in humans and animal models[67]. Therefore, while the reasons for the down-regulation are unknown, the synaptic gene down-regulation in tau-negative neurons may represent an evolutionarily conserved aging-related phenomenon which is exacerbated in AD.

One limitation of this study is the profiling of a single cell population. This study was designed to specifically evaluate changes in gene expression associated with intraneuronal tau. The hippocampus was selected as a region heavily affected by tau pathology in PART and AD and, as this is not a single cell technique, this region was also used because of the ability to select a relatively uniform excitatory neuronal population based on anatomy and morphology. While the gene expression changes were similar in PART and AD in excitatory neurons, other cell types such as inhibitory interneurons, astrocytes, and microglia were not assessed. Microglia and astrocytes show differential gene expression in AD[68-71]. Though mRNA expression has not been examined in PART in glia, rare studies of neuroinflammation in PART do not show overt neuroinflammation[70]. It is possible, even likely, that disease-specific transcriptional signatures differentiating AD from PART would be identified in other cells types. Therefore, this study does not provide evidence that PART and AD are the same disease process. However, the similarities in tau-associated transcriptional regulation would raise the possibility of a similar pathogenesis in the hippocampal tau pathology.

This study is also limited in the conclusions that can be drawn between disease and control groups due to control mRNA variability and the number of control samples. The measurements of gene changes associated with tau pathology had greater statistical power by comparing groups within the same sample (tau-positive versus tau-negative). Few DEGs were identified in AD, confirming that this study was not sufficiently powered to detect gene changes in disease versus control comparisons. The DEGs identified in PART overlapped significantly with those altered in AD and may reflect a signature of aging rather than a specific signature associated with PART. Future studies with more samples would be required to determine if there are gene signatures in the brain specific to PART.

## Conclusions

In this study, we identify novel gene expression signatures and synaptic gene changes associated with intraneuronal tau pathology in primary age-related tauopathy (PART) and Alzheimer’s disease (AD). To our knowledge, this is the first study of gene expression changes associated with tau pathology in PART. The gene expression changes associated with intraneuronal tau pathology were similar in PART and AD, raising the possibility that the two tau aggregates may be mechanistically related. This study also identified that decreased synaptic gene expression in neurons in PART and AD was largely driven by neurons lacking intraneuronal tau pathology. Synaptic gene expression in tau-positive neurons was relatively preserved compared to controls or increased compared to tau-negative disease neurons. These tau-associated gene expression changes were confirmed in a published transcriptional dataset of cortical neurons with tau pathology in AD. The novel molecular signatures identified in this study provide insight into previously underappreciated transcriptional changes in neurons with tau pathology in PART and AD, and highlight the importance of accounting for pathology in molecular studies in neurodegeneration.

## Data Availability

The code used in the current study is available in Github, https://github.com/SteinOBrienLab/AD_PART_Tau_23. The datasets generated and analyzed during the current study will be available at the Neuroscience Multi-omic Data Archive (NeMO) upon publication of the report.

## Declarations

### Ethics Approval and Consent to Participate

All work in this study was approved by the Johns Hopkins Institutional Review Board. All tissue used in this study was donated to the Johns Hopkins Brain Resource Center with consent from next of kin.

### Authors Contributions

MM, JCT, RLH, and GLSO designed the study. MM, JRO, JCT, WRB, and OP participated in tissue selection and pathologic analysis. MM, HG, and EMM developed experimental protocols and performed data collection. MM, RP, and GLSO performed data analysis. MM and GLSO wrote the manuscript. All authors edited and approved the manuscript.

## Acknowledgements

We would like to acknowledge Prajan Divakar, Nanostring, for technical support. The macro code (Suppl. File 1) was provided courtesy of Research and Development at Nanostring (Seattle, WA).

## Additional Material

Supplemental_Figures (.pdf)

**Supplemental Figures 1-6**

Additional_File_1 (.txt)

**Image J Macro for GeoMx Neuron Selection**

Code provided for manually generating Image J masks of neurons with and without tau pathology, courtesy of Nanostring Research and Development.

Additional_File_2 (.xlsx)

**Gene Expression Analysis in Tau Positive versus Tau Negative Neurons in AD and PART** Differentially expressed genes were defined as a log2 fold change >0.2 and false discovery rate-corrected p value of <0.05.

Additional_File_3 (.xlsx)

**Gene Expression Analysis in Disease versus Control**

Differentially expressed genes were defined as a log2 fold change >0.2 and false discovery rate-corrected p value of <0.05.

Additional_File_4 (.xlsx)

**Fold Change in SynGO Annotated Genes**

Quantified genes were queries through SynGO (https://www.syngoportal.org/) to identify a total of 353 genes with synaptic annotations.

Additional_File_5 (.xlsx)

**PatternMarkers Analysis**

PatternMarkers analysis was performed after coordinated gene association in pattern sets (Co-GAPS) analysis to determine the genes significantly associated with each pattern. This analysis results in a score and rank for each gene, and a list of genes significantly associated with that coordinated gene expression pattern.

Additional_File_6 (.xlsx)

**ShinyGO Analysis of Tau Up Pattern**

The genes significantly associated with the Tau Up pattern in the PatternMarkers analysis were submitted for gene ontology analysis using the “GO Biological Process” pathway set in ShinyGO (version 0.77). The list of all gene quantified in hippocampal neurons was used as the background gene set. The top 20 pathways are listed.

