## Supplemental Figures for "Transcriptional Signatures of Hippocampal Tau Pathology in Primary Age-Related Tauopathy and Alzheimer’s Disease"

**Supplemental Figures 1-6**

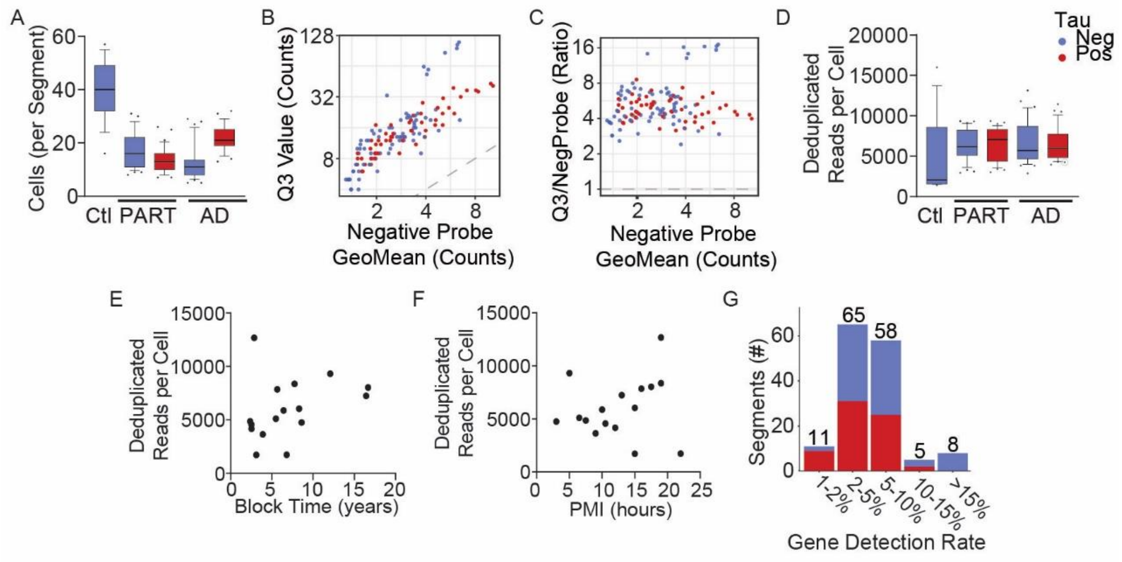


**Supplemental Figure 1. mRNA Collection and Quality Analysis**. (A) Number of cells collected per segment by disease group and tau status. (B) Third quartile (Q3) versus negative control probe counts and (C) the ratio of the third quartile counts over the negative control probe counts showing good quality mRNA signal. (D) Similar deduplicated mRNA counts per cell among all groups by disease group and tau status. (E,F) No association of tissue time in block (E, *p*=0.22) or post-mortem interval (F, *p*=0.58) with the number of deduplicated mRNA reads per cell by simple linear regression analysis. (G) Gene detection rate (percent of whole transcriptome atlas) was between 2-10% for most groups/segments collected. Segments with less than 2% gene detection were excluded from further analysis. AD, Alzheimer's disease; Ctl, control; Neg, tau pathology negative; PART, primary age-related tauopathy; Pos, tau pathology positive; Q3, third quartile value. A,D – Error bars are 10-90$^{th}$ percentile.

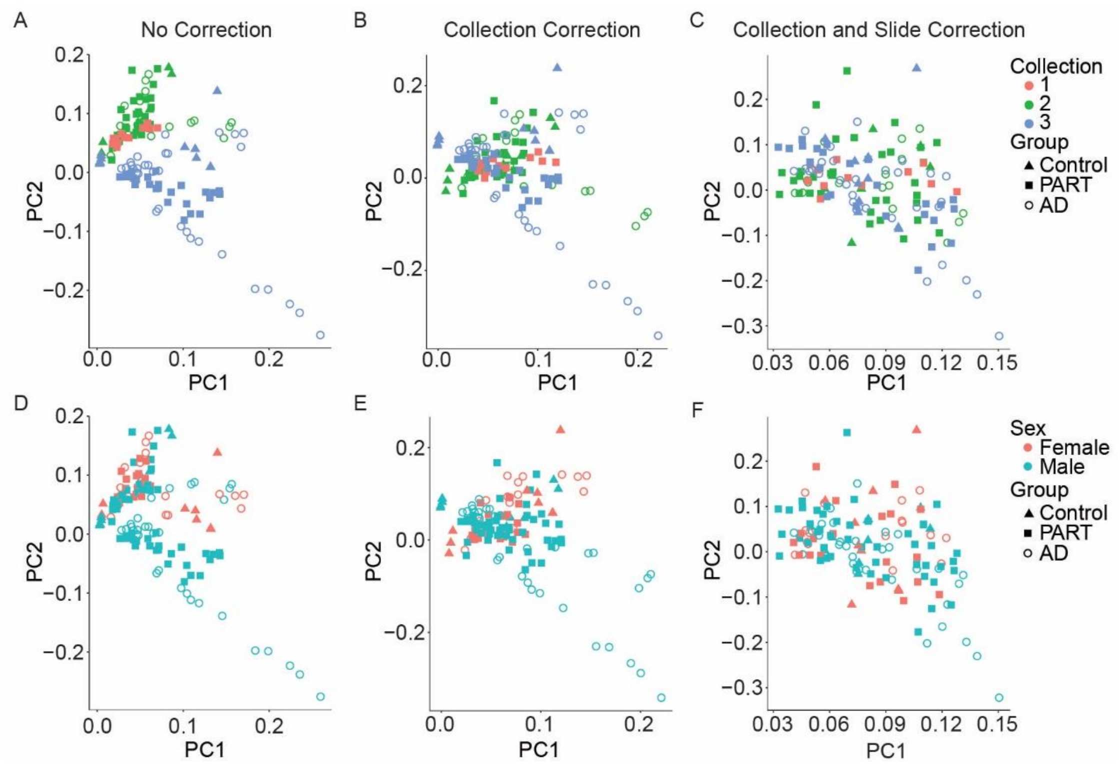


**Supplemental Figure 2. Principal Components Analysis with Collection Day and Slide Correction**. Principal components analysis was conducted to assess variability associated with collection day, disease, and sex. Collection day was the primary driver of data variability in the raw transcriptional data (A), which was eliminated after ComBat batch correction using either the collection day alone (B) or collection day and slide (C) as the batch variable. Sex was not a large driver of transcriptional variability without ComBat batch correction (D), or with ComBat correction using either collection day (E) or collection day and slide (F) as the batch variables. AD, Alzheimer's disease; PART, primary age-related tauopathy; PC, principal component.

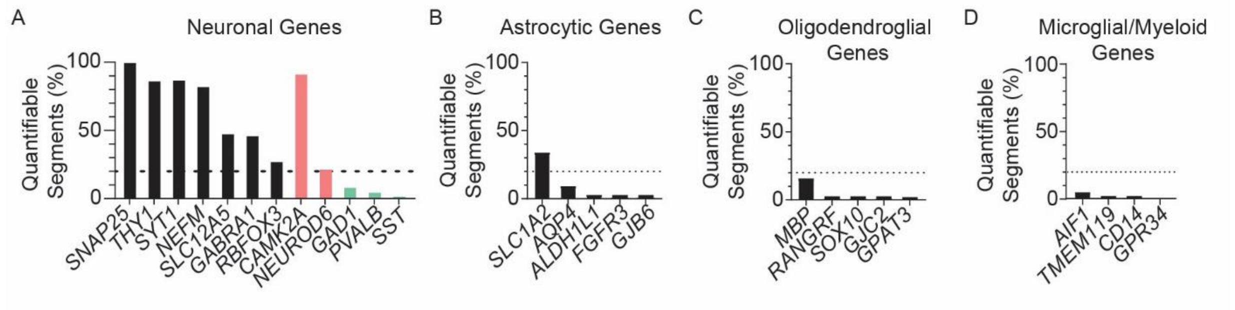


**Supplemental Figure 3. Neuronal Gene Enrichment in Quantified mRNA**. Genes were included in the quantitative analysis if they were greater than 2 standard deviations above the GeoMean of the negative control probes in more than 20% of segments (dotted line). (A) Neuron-expressed genes were frequently detected in the segments, including genes specific to excitatory neurons (peach, *CAMK2A*, *NEUROD6*). Genes specific to inhibitory interneurons were not reliably detected in most segments (green, *GAD1*, *PVALB*, *SST*). (B) Of the astrocytic genes, only one (*SLC1A2*) was quantifiable, however, this gene can show lower level expression in neurons. (C,D) Oligodendroglial and Microglial/myeloid lineage genes were infrequently detected.

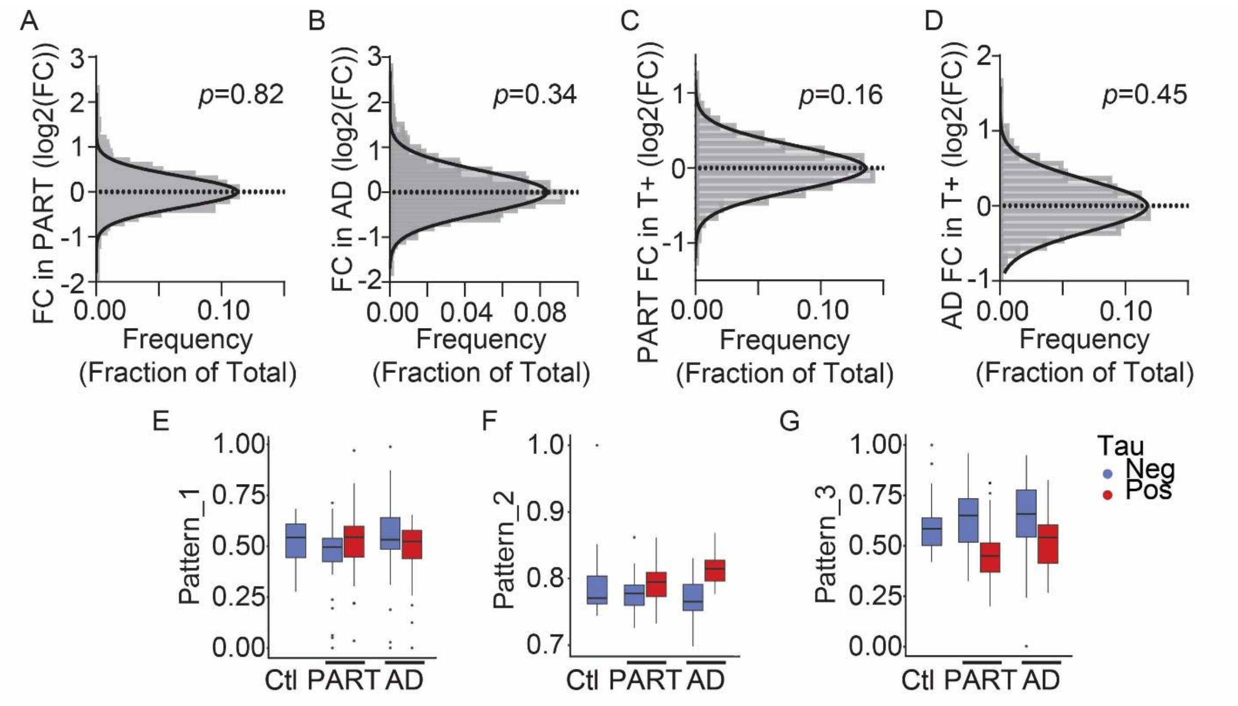


**Supplemental Figure 4. Gene Expression Analysis Not Associated with Intraneuronal Tau Pathology**. (A-D) The distribution of the log2 fold change of all quantified genes for the following comparisons (A) PART versus control, (B) AD versus control, (C) tau-positive versus tau-negative in PART, (D) tau-positive versus tau-negative in AD. The curves were generated by Gaussian least squares fit and the mean is not different from 0 by extra sum of squares F test; dotted line at 0. (E-G) CoGAPS patterns not strikingly associated with disease group or intraneuronal tau. AD, Alzheimer's disease; Ctl, control; FC, fold change; Neg, tau pathology negative; PART, primary age-related tauopathy; Pos, tau pathology positive; T-, tau pathology negative; T+, tau pathology positive. E-G - Error bars are 95% CI.

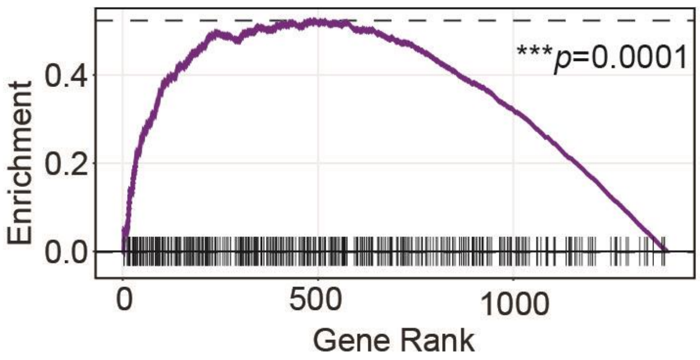


**Supplemental Figure 5. SynGO Annotated Genes Enriched in Tau Up Pattern**. The genes significantly associated with the Tau Up pattern after PatternMarkers analysis were analyzed for enrichment of synaptic genes (SynGO annotated) by fast gene set enrichment analysis. All quantified genes are ordered by rank on the x-axis with genes annotated in SynGO marked with a line.

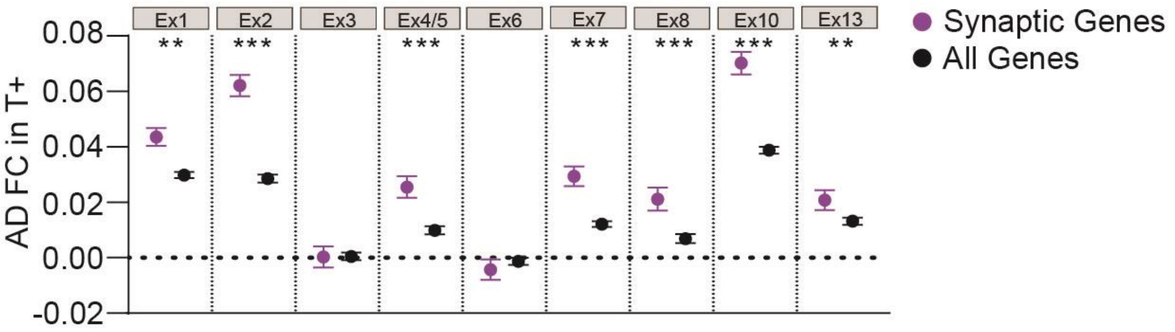


**Supplemental Figure 6. Additional Synaptic Gene Expression Patterns Associated with Cortical Intraneuronal Tau in Single Cell Dataset**. Publicly available single soma RNA-seq data from cortical neurons in AD [22] was analyzed for synaptic gene expression changes. Excitatory neuronal grouping was performed as in the published analysis for tau positive and negative neurons. Mean expression values showing increased synaptic gene expression (SynGO annotated) compared to expression of all quantified genes in each excitatory neuronal group. Significance is comparing mean of Gaussian least squares fit curve of synaptic genes and all measured genes by extra sum of squares F test. AD, Alzheimer's disease; FC, log2 fold change; T+, tau-positive. Error bars are SEM.
