## Additional File 1 for "Transcriptional Signatures of Hippocampal Tau Pathology in Primary Age-Related Tauopathy and Alzheimer’s Disease"

colorArray = newArray("Blue","Green","Yellow","Red");	//this array has the color channels in order

height = getHeight;
width = getWidth;

name = getInfo("image.filename");
namelength =lengthOf(name);
name = substring(name,0,namelength-5);		//this clips the name to just ROI###


run("Make Composite", "display=Composite");	//makes tiff into a composite


//moves through the channels of the image, up to number of chanels. sets colors to match ColorArray.
for (i=0; i<nSlices; i++){		
	Stack.setChannel(i+1);
	run(colorArray[i]);
	run("Enhance Contrast", "saturated=0.35");
	}
	
setForegroundColor(255, 255, 255);


roiManager("deselect");
